# The living conditions and experiences of caregivers: a cross-sectional study in public hospitals of Yaoundé-Cameroon

**DOI:** 10.1101/2025.09.07.25335277

**Authors:** Rick Tchamani, Richard Tchadpa, Constantine Tanywe Asahngwa, Leonelle Ariane Fengaing Sikati, Fabrice Zobel Lekeumo Cheuyem

## Abstract

**Background:** In Cameroon’s overburdened healthcare system, informal caregivers are partners in patient care. This study aimed to address the existing knowledge gap by providing a description of the living conditions, challenges, and interactions of caregivers in Yaoundé’s public hospitals.

**Methods:** A cross-sectional study was conducted from April to July 2025 at two public hospitals in Yaoundé. Participants included individuals aged 21 or older who had cared for a hospitalized patient for at least two days. Data were collected through face-to-face interviews using a pre-tested questionnaire. The questionnaire gathered information on socio-demographics, hospital experiences, patient care details, and relationships with medical personnel. Data were analyzed with R Statistics, and binary logistic regression was used to identify factors associated with altercations between caregivers and health workers.

**Results:** A total of 327 caregivers participated in the study. The majority were women (85.0%), and a significant proportion were aged between 41 and 50 years (29.4%). Most caregivers were married (58.7%) and had attained a secondary education level (47.7%). More than half of the caregivers (51.1%) had spent between 4 to 7 days in the hospital, during which they faced dire living conditions: 37.9% slept on benches in hallways, and 30.9% slept on pieces of cloth on the floor. Consequently, 51.7% reported never getting proper sleep, primarily due to poor sleeping conditions (90%) and the presence of mosquitoes (77%). Regarding their role in patient care, 73.7% performed various caregiving tasks, including cleaning patients (69%), dressing patients (61%), and feeding patients (56%). Additionally, 29.4% reported experiencing verbal altercations with healthcare providers, mainly due to patient abandonment (51%), neglect (34%), poor communication (35%), and delays in medical services (29%). Multivariate analysis revealed that caregivers who sometimes experienced difficulties sleeping (adjusted Odds Ratio (AOR) = 4.02; 95% CI: 1.27-17.8; *p* = 0.033) and those who knew their relative’s diagnosis (AOR = 3.23; 95% CI: 1.65-6.86; *p* = 0.001) were significantly more likely to engage in verbal confrontations with healthcare staff.

**Conclusion:** The findings show that informal caregivers in Yaoundé’s public hospitals operate under conditions that compromise their physical and psychological well-being. The lack of adequate sleeping arrangements and caregiving burden contributes to tensions. The link between sleep deprivation, knowledge of the patient’s diagnosis, and altercations with health workers highlights an urgent need for systemic interventions. Addressing these issues requires clear policies that formally recognize the role of caregivers, ensure their basic needs are met, and foster collaborative relationships.

## Introduction

Caregivers in hospitals are typically patient companions who assume various roles, such as assistant, representative, navigator, or planner [1]. They are most often family members or close relatives of the patient who take on this responsibility [2,3].

In Cameroon, hospitals are facing high demand for care and an overcapacity of hospital beds due to a significant influx of patients [4,5]. However, the country suffers from a shortage of healthcare personnel compared to predefined standards [4,6]. In this context, caregivers are seen as indispensable partners on whom the healthcare system relies. Their presence and actions, which may at times lead to conflict with healthcare staff, are a necessary and complementary part of the care and services dedicated to the patient’s comprehensive well-being. Their interventions usually include ensuring patients are fed, assisting with hygiene, dressing them, monitoring their condition, preparing their breakfast, reminding them of medication times, taking their temperature, and even going to pharmacies to purchase prescribed medications [5,7].

However, their stay in hospital facilities takes place under precarious conditions, with nights often spent in difficult circumstances [7]. On a daily basis, patient caregivers face numerous challenges, including physical and psychological strain, financial difficulties, social isolation, and a lack of support [8]. Although their involvement is increasingly prominent in healthcare systems in developing countries [9–11], these caregivers generally lack any specific skills for this task [7]. Moreover, the hospital environment is experiencing a surge in violence, influenced by interactive factors such as staff behavior, patient behavior, hospital conditions, professional roles, and waiting times [12]. This situation negatively impacts patient care and leads to demotivation and disengagement among healthcare professionals. Among patients, it exacerbates anxiety and stress, thereby compromising treatment effectiveness [13].

Despite their integral role in patient care, the daily realities and experiences of caregivers in Cameroonian hospitals remain insufficiently documented. This critical knowledge gap hinders the development of a supportive and effective healthcare ecosystem. Therefore, this study is critically needed to provide a detailed description of their living conditions in healthcare settings in Yaoundé. The evidence-based data generated from this research will serve as a vital resource for hospital managers and policymakers, enabling them to develop and implement health policies that improve the well-being of this essential population and, ultimately, enhance the quality of patient care.

## Methods

### Study design & period

A cross-sectional study with descriptive and analytical aims was conducted in two public reference hospitals located Yaoundé (Biyem-Assi District Hospital and Military Hospital of Region No.1). The study was carried out over a three-month period, from April to July 2025.

### Setting

Yaoundé, the political capital of Cameroon’s Centre Region, has an estimated population of 1.5 million, representing all of the country’s ethnic groups. Its demographic structure is characterized by a very young population and a gender imbalance favoring men. Between 70% and 80% of the urban population resides in spontaneous neighborhoods [14]. The Biyem-Assi Hospital, a fourth-category, first-level referral hospital, is located in the Yaoundé VI subdivision. The Military Hospital of Region No. 1, a first-category, third-level referral hospital, is situated in the Yaoundé III subdivision [15]. The Both hospitals cumulated in 2024, approximately 73,000 consultations, 300 beds, 9,900 hospitalizations, and provided more than 35,000 days of hospitalizations [16].

### Study participants & selection criteria

Any individual at least 21 years old who was caring for a patient hospitalized in one of the selected facilities for 48 hours or more was eligible. All caregivers meeting these criteria and who provided an informed consent were included in the study.

### Sampling method

The sample size was calculated using the single proportion formula (n = [Zα / 2] 2P (1-P) / d2) at 95% confidence interval, where Zα / 2 = 1.96, P = 50% prevalence since there was no similar study conducted previously in the study area and d = 5% of marginal. Using the above formula the estimated sample size of 384 was obtained. Caregivers were approached at their various departments and asked to participate. A convenience non-probabilistic technique was used to enroll consenting caregivers.

### Data collection tool and procedure

The study tool was a questionnaire designed to collect Sociodemographic, Hospital stay experience (sleeping location, tasks performed, awareness of patient care details), and Relationships with medical personnel. the questionnaire was pre-tested and then administered through face-to-face interview method with consenting caregivers.

### Data processing and analysis

Data were entered, exported, recoded as necessary, and analyzed using R Statistics version 4.4.2 [17]. The respondent characteristics were presented as counts and frequencies. The predictor consisted of sociodemographic characteristics including age, gender, study level, living situation, marital status, professional status, and monthly income. The dependent variable was the experience of an altercation with health worker. Simple and multiple binary logistic regressions were used to assess the strength of association between variables. The selection of predictors that best fit the model was done stepwise using the Akaike Information Criterion [18]. The model with the lowest index was then selected. A *p*-value <0.05 was considered statistically significant.

## Results

### Sociodemographic characteristic of participants

A total of 327 caregivers were recruited. The majority were women (85.0%), with a significant portion aged between 41 and 50 years (29.4%). Most caregivers were married (58.7%) and had attained a secondary education level (47.7%). In terms of employment, the majority were self-employed (45.3%), with a monthly income typically between 50,000 and 100,000 CFA francs (28.1%) (Table 1).

**Table 1.** Sociodemographic characteristics of study participants, Yaoundé, 2025, Cameroon (n = 327)

| Characteristics | Count (n) | Frequency (%) |
| --- | --- | --- |
| **Age** |  |  |
| 21 – 30 | 55 | 16.8 |
| 31 – 40 | 73 | 22.3 |
| 41 – 50 | 96 | 29.4 |
| 51 – 60 | 81 | 24.8 |
| 61+ | 22 | 6.7 |
| **Sex** |  |  |
| Female | 278 | 85.0 |
| Male | 49 | 15.0 |
| **Study level** |  |  |
| No formal education | 33 | 10.1 |
| Primary | 85 | 26.0 |
| Secondary | 156 | 47.7 |
| Tertiary | 53 | 16.2 |
| **Marital status** |  |  |
| Single | 99 | 30.3 |
| Divorced | 6 | 1.8 |
| Married | 192 | 58.7 |
| Widow(er) | 30 | 9.2 |
| **Professional status** |  |  |
| Unemployed | 98 | 30.0 |
| Private sector | 54 | 16.5 |
| Public sector | 27 | 8.3 |
| Self-employed | 148 | 45.3 |
| **Income (in CFA Francs)** |  |  |
| None | 98 | 30.0 |
| □ 50000 | 69 | 21.1 |
| 50000-100000 | 92 | 28.1 |
| > 100000 | 68 | 20.8 |

More than half of the caregivers (51.1%) had already spent between 4 to 7 days in the hospital. Their sleeping conditions were often dire; most slept on benches in hallways (37.9%) or on pieces of cloth on the floor (30.9%). Consequently, over half (51.7%) reported they were never able to get proper sleep (Table 2). The primary reasons for their insomnia were poor sleeping conditions/environment (90%) and the presence of mosquitoes (77%) (Fig. 1).

**Table 2.** Caregiver experience in hospital settings, Yaoundé, 2025, Cameroon (n = 327)

| Variable | Count(n) | Frequency (%) |
| --- | --- | --- |
| **Current period of caregiving (in days)** |  |  |
| 2-3 | 87 | 26.6 |
| 4-7 | 167 | 51.1 |
| 8+ | 73 | 22.3 |
| **Patient care planning known** |  |  |
| No | 200 | 61.2 |
| Yes | 127 | 38.8 |
| **Patient diagnosis known** |  |  |
| Under investigation | 30 | 9.2 |
| No | 88 | 26.9 |
| Yes | 209 | 63.9 |
| **Patient oral medication schedule known** |  |  |
| No | 74 | 22.6 |
| Yes | 169 | 51.7 |
| Non applicable | 84 | 25.7 |
| **Sleeping site** |  |  |
| Outdoor in the open air | 51 | 15.6 |
| At the patient's bedside | 27 | 8.3 |
| In my car | 4 | 1.2 |
| Same bed as the patient | 20 | 6.1 |
| On the bench | 124 | 37.9 |
| On a scarf on the floor | 101 | 30.9 |
| **Easy to fall asleep during the night** |  |  |
| Yes always | 27 | 8.3 |
| Sometimes | 131 | 40.1 |
| Never | 169 | 51.7 |
| **Carried out in-hospital task** |  |  |
| No | 86 | 26.3 |
| Yes | 241 | 73.7 |
| **Experienced an altercation/verbal violence with health worker** |  |  |
| No | 231 | 70.6 |
| Yes | 96 | 29.4 |

**Fig. 1.**
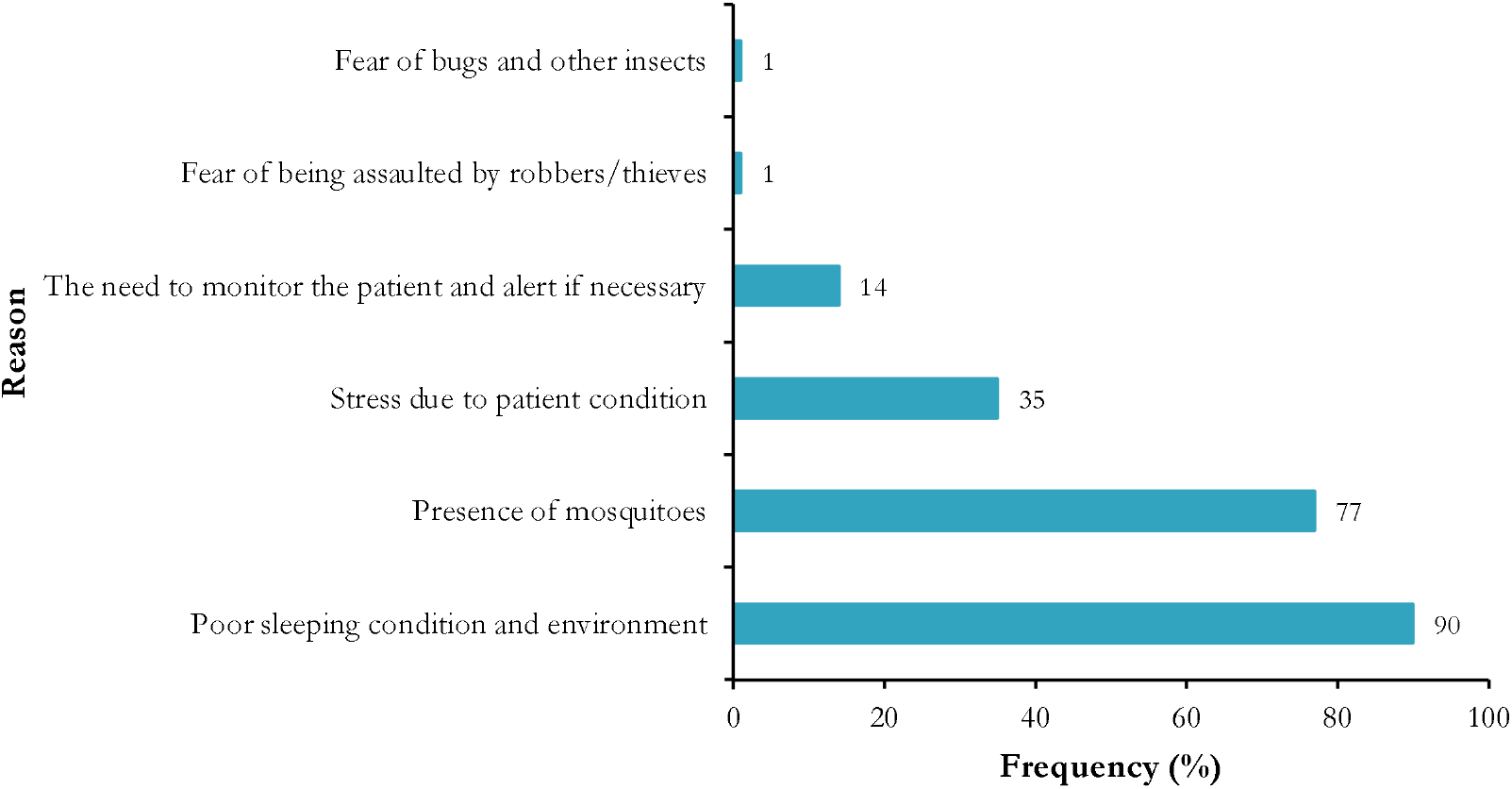
Reasons for disturbed sleep among caregivers in health facilities, Yaoundé, 2025, Cameroon (n = 300)

Regarding their knowledge of patient care, 127 out of 327 participants (38.8%) were aware of their relative’s care schedule. However, a much lower proportion (26.9%) did not even know the illness their patient was suffering from (Table 2).

Several participants reported experiencing verbal violence with healthcare providers. The main reasons cited for these incidents included patient abandonment (51%), patient neglect (34%), poor communication (35%), and delays in medical services (29%) (Fig. 2).

**Fig. 2.**
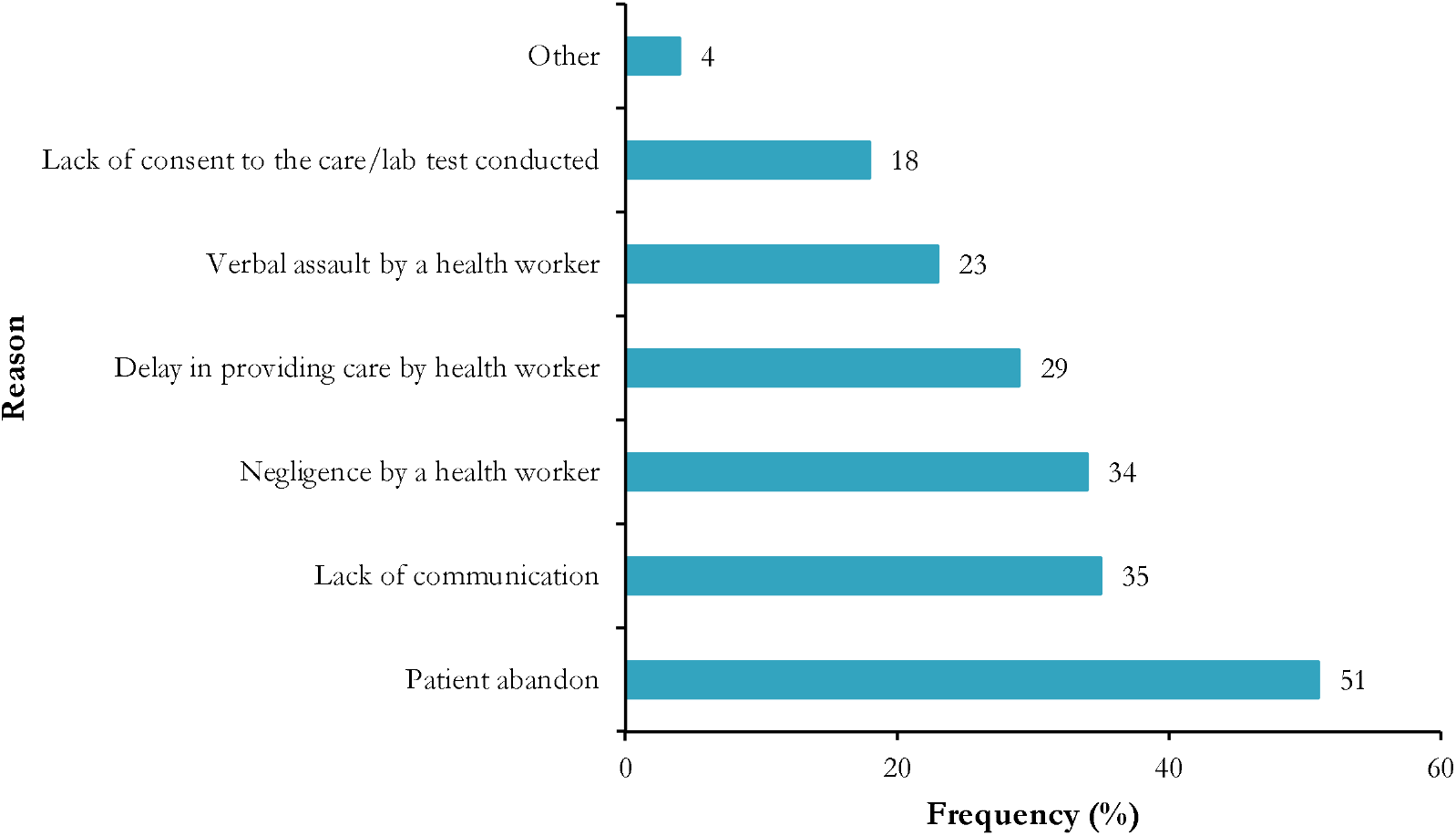
Reported instigator of hospital altercation experienced by patients’ caregivers in Yaoundé health facilities, 2025, Cameroon (n = 96)

Most of participants (73.7%) reported performing various caregiving tasks. Their primary duties involved cleaning patients (69%), dressing patients (61%), feeding patients (56%), administering medication (28%), and making patient beds (13%) (Fig. 3).

**Fig. 3.**
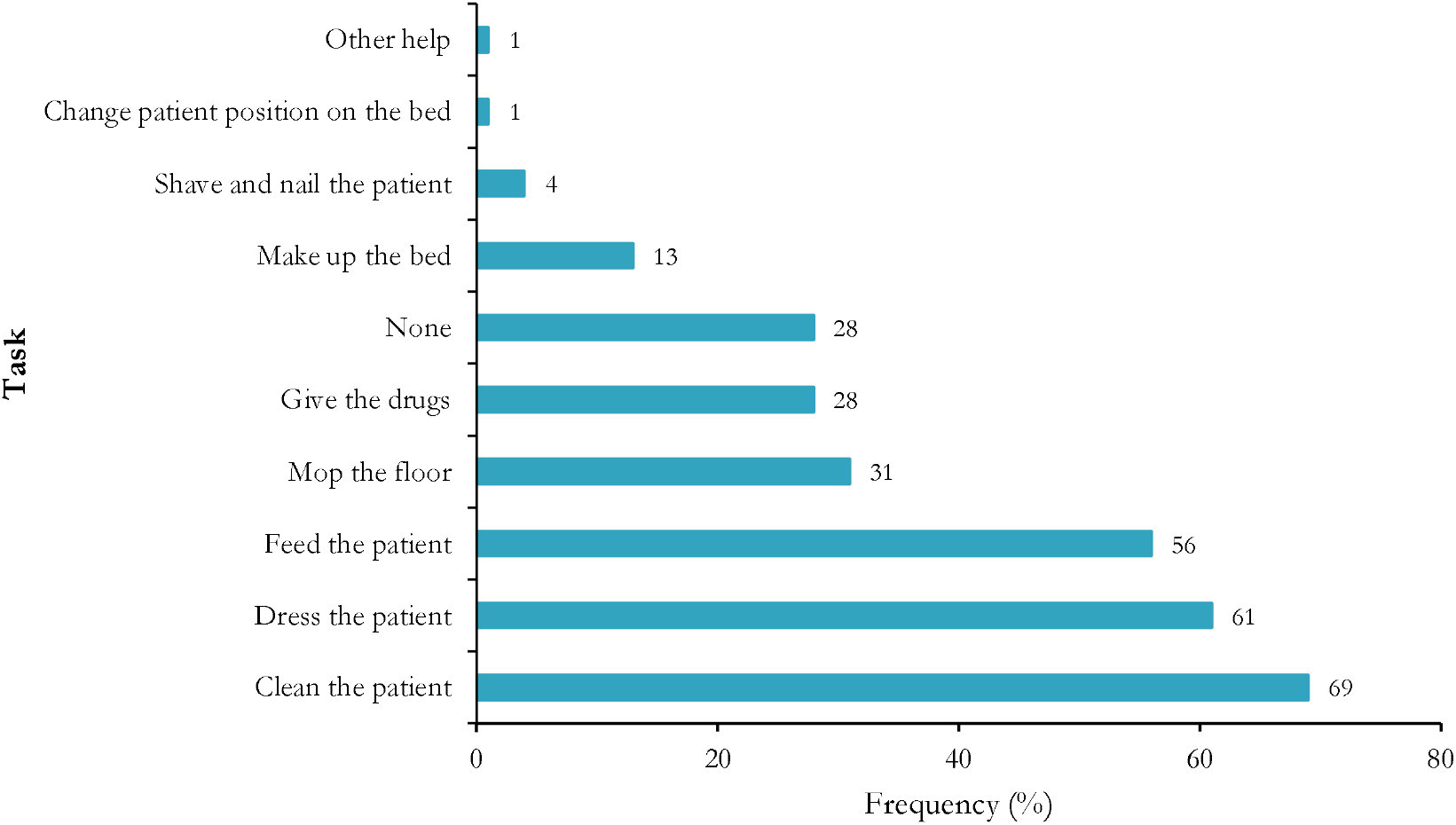
Tasks carried out within health facilities by caregivers, Yaoundé, 2025, Cameroon (n = 241)

### Predictors of the risk of altercation between caregivers and health workers

Risk of experiencing an altercation with a healthcare provider was: 96/327: 29.4% (95% CI: 24.5-34.6). The multivariate analysis shows a significant association between marital status, knowledge of the patient’s diagnosis and the sleeping experience of caregivers. Caregivers who sometimes experience difficulties in sleeping (AOR = 4.02; 95% CI: 1.27-17.8; *p* = 0.033) or who knew their relative’s diagnosis (AOR = 3.23; 95% CI: 1.65-6.86; *p* = 0.001) were four and three times respectively more likely to engage in verbal confrontations with healthcare providers (Table 3).

**Table 3.**
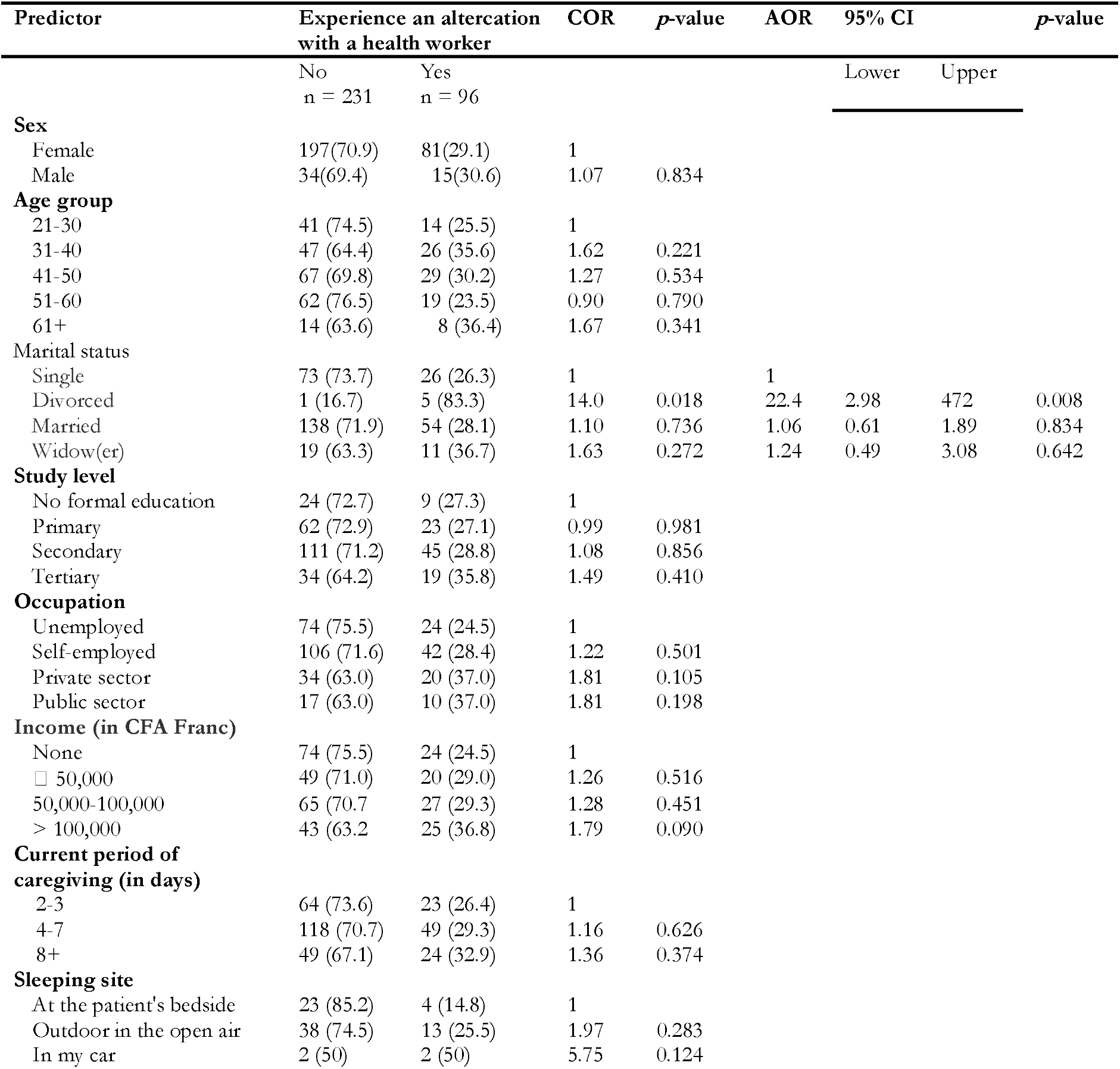
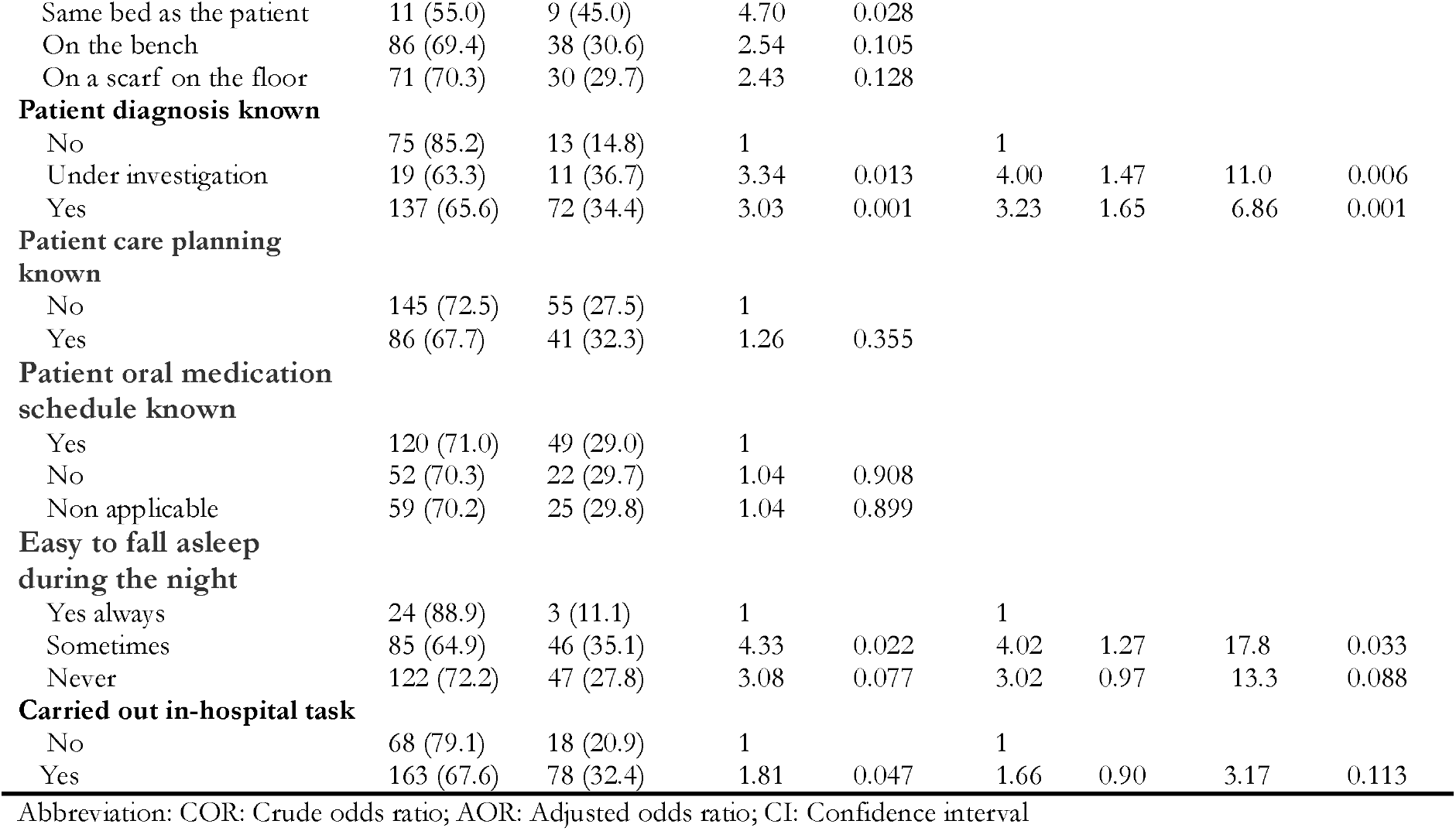
Simple and multiple logistic regression of predictor of altercation between cargivers and health worker in hospital setting, Yaoundé, 2025, Cameroon (n = 327)

## Discussion

This study aimed to document the daily living conditions of caregivers in two referral hospitals in Yaoundé. The findings reveal that their hospital stay is characterized by significant hardships and frequently conflictual relationships with healthcare staff.

Most of the participants were women over 41 years old, a finding consistent with observation in a study conducted in UK [19]. This trend may reflect women’s increased likelihood of assuming maternal caregiving roles with advancing age, making them more inclined to care for ill family members [2,3]. Half of the caregivers were either unemployed or self-employed, with monthly incomes below 50,000 CFA francs. This pattern likely reflects hospital care demands, as families typically designate the most available member to provide prolonged patient support [20].

Regarding their hospital experience, over a quarter of caregivers performed caregiving tasks for their relatives, primarily including feeding, bathing, dressing, and administering medication. A study conducted in Cameroon highlighted the involvement of caregivers in clinical care [5]. Although these tasks typically fall under healthcare staff responsibilities, overburdened providers, despite being aware of their duties, often fail to perform them due to staffing shortages [4,21]. Consequently, caregivers are compelled to take on these roles to ensure proper treatment adherence.

Over 50% of caregivers slept outdoors or on hallway benches. This situation stems from the lack of clear regulatory frameworks defining their status in Cameroonian healthcare facilities [5,20], exposing them to precarious conditions that may increase disease susceptibility [22]. Confronted with this challenge, health authorities must urgently develop policies that specifically address patient attendants’ needs within the healthcare system. Therefore, it is evident that, exposed to these living conditions in healthcare facilities, caregivers may experience the onset of healthcare associated infections such as malaria, diarrheal diseases, stress, hypertension, and even unexplained deaths. This situation indeed raises the question of how to properly consider and care for caregivers as partners in the healthcare system.

Some caregivers reported sharing beds with patients, despite this practice being strictly prohibited, even prohibiting sitting on patient beds [23]. This concerning practice highlights the urgent need to implement Behavior Change Communication sessions to educate caregivers about associated risks and to modify these unsafe behaviors [24].

In the Cameroonian context, it is entirely understandable for these care partners to offer a critical judgment on healthcare services. While it is true that the caregiver is a potential patient due to the uncertainty surrounding the outcome of their sick loved one, this suffering must still be taken into account. A question, an inquisitive look, a manifestation of dissatisfaction, or even violence may be signs and symptoms associated with this distress.

The study showed that caregivers experience difficult living conditions in healthcare facilities. They often dedicate all of their time to caring for their sick loved ones, to the point of neglecting themselves. They also lack a framework that could offer them a minimum level of security. Some sleep on the floor, while others stay awake simply to remain as close as possible to the patient.

These living conditions have serious physical and psychological consequences, including fatigue, stress, and body aches. The precariousness of their situation also exacerbates these issues due to the uncertainty regarding the patient’s prognosis [25–27]. In most African healthcare facilities, a caregiver’s suffering is intensified when they are financially incompetent or when the hospitalization is prolonged. This leads to a strong feeling of shame, social isolation, and a high tendency toward depression or suicide [25].

Nights spent outdoors or in makeshift shelters are a genuine ordeal for caregivers, who are exposed to cold and various weather conditions. A study conducted in Cameroon revealed that a caregiver’s suffering severely affects the sick person [28]. Poor hospital hygiene conditions also lead to the proliferation of mosquitoes, and it is not uncommon for some caregivers to contract severe forms of malaria.

Thus, the deprivation of sleep, adequate housing, and exposure to diseases in the hospital environment are all factors that limit the caregivers’ satisfaction with the care they provide, even when quality care and services have been offered to their sick loved ones [26,27].

Regarding the causes of altercations between caregivers and healthcare staff, communication breakdowns represent a leading cause of verbal aggression toward healthcare workers. These findings align with observations documented in Cameroon [29]. Yet information access remains a fundamental right for caregivers, underscoring the critical need to train clinical staff in ethical engagement with caregivers, as stipulated in patient attendant charters [23].

Patient abandonment or neglect was identified as a key driver of verbal violence between healthcare workers and caregivers. However, contrasting findings emerged from a seven-country European study [30]. In our setting, this discrepancy may reflect staffing shortages and ward overcrowding, which severely limit the time available for individual patient care [31].

Multivariate analysis revealed that sleep deprivation tripled the risk of verbal aggression toward healthcare workers, a finding consistent with prior observational studies [12,30]. This aligns with established evidence demonstrating that sleep deficiency impairs emotional regulation and increases irritability [32,33].

Knowledge of the patient’s diagnosis tripled the risk of verbal aggression toward healthcare staff, consistent with findings reported in Israel [12]. This association may stem from stress, depression, anxiety, or the fear of losing a loved one [34]. Furthermore, socioeconomic factors prevalent in our setting, including widespread financial insecurity, a lack of universal health coverage, and low health insurance enrollment, exacerbate tensions, resulting in targeted hostility toward. medical personnel [35].

## Strengths and limitations

This study provides quantitative evidence of the precarious living conditions faced by caregivers in Cameroonian public hospitals and identifies specific, modifiable drivers of conflict. It highlights a critical communication gap and offers evidence for policy intervention by directly linking systemic issues to outcomes that affect hospital safety and care quality. The findings provide a clear mandate for developing caregiver charters and support systems. However, this study bears several limitations. This study’s use of a cross-sectional design limits the ability to establish causal relationships between the observed factors and outcomes. The reliance on self-reported data through interviews introduces the potential for recall and social desirability bias among participants. Furthermore, the non-probabilistic convenience sampling method employed at two hospitals in a single city might restrict the generalizability of the findings to the broader population of caregivers across Cameroon. Therefore, future research should employ longitudinal or mixed-methods designs to better establish causality between caregiver hardships and health outcomes, while also capturing the nuanced emotional and experiential aspects of their role. Expanding the geographical scope to include rural health facilities and a more diverse range of hospitals would enhance the generalizability of the findings and allow for regional comparisons.

## Conclusions

This study revealed that informal caregivers in Yaoundé’s public hospitals are an indispensable yet severely vulnerable population, whose critical role in patient care is undermined by dire living conditions, including sleep deprivation due to unsafe and inadequate sleeping arrangements, a high burden of performing clinical tasks, and a significant prevalence of conflict with healthcare staff, which is predicted by their psychological distress and physical exhaustion. Addressing these issues requires the development of clear policies that formally recognize the role of caregivers, ensure their basic needs are met, and foster collaborative relationships with healthcare providers. By improving the conditions and support systems for caregivers, hospitals can not only enhance their well-being but also promote a more effective and compassionate healthcare environment, ultimately benefiting both caregivers and patients.

## Data Availability

All data generated or analyzed during this study are included in this published article.

## Abbreviations

aOR: Adjusted Odds Ration
CI: Confidence Interval
cOR: Crude Odds Ratio

## Declarations

### Authors’ Contribution

- Conceptualization: R.T.1 and F.Z.L.C.;
- Data curation: R.T.1 and F.Z.L.C.;
- Formal analysis: R.T.1 and F.Z.L.C.;
- Investigation: R.T.1 and F.Z.L.C.;
- Methodology: R.T.1, R.T.2, C.T.A., L.A.F.S. and F.Z.L.C.;
- Software: R.T.1 and F.Z.L.C.;
- Supervision: F.Z.L.C.;
- Visualization: R.T.1, R.T.2, C.T.A., L.A.F.S. and F.Z.L.C.;
- Writing – original draft: R.T.1 and F.Z.L.C
- Writing – review and editing: R.T.1, R.T.2, C.T.A., L.A.F.S. and F.Z.L.C.;
- Validation: R.T.1, R.T.2, C.T.A., L.A.F.S. and F.Z.L.C.;

### Ethical approval statement

This protocol was approved by the Human Health Research Ethical Review Committee for the Centre Region (CRERSH - Ce) and the ethical clearance: CE Nº 00379/CRERSH/2025 issued. Informed consent was obtained from participants prior to inclusion in the study. All methods were performed in accordance with declaration of Helsinki.

### Consent for publication

Not applicable.

### Availability of data and materials

All data generated or analyzed during this study are included in this published article.

### Competing interests

The author declares no conflict of interest and have approved the final version of the article.

### Funding source

This research did not receive any specific grant from funding agencies in the public, commercial or not-for-profit sectors.

## Acknowledgements

Our gratitude goes to the health staff who agreed to participate in this study and to the manager of the health facility who gave an authorization to conduct the study.

## Notes

### Competing Interest Statement

The authors have declared no competing interest.

### Author Declarations

Human Health Research Ethical Review Committee for the Centre Region (CRERSH - Ce)

